# Population simulations of COVID-19 outbreaks provide tools for risk assessment and continuity planning

**DOI:** 10.1101/2020.04.13.20064253

**Authors:** Bo Peng, Christopher I. Amos

## Abstract

**Objectives:** We developed COVID-19 Outbreak Simulator (https://ictr.github.io/covid19-outbreak-simulator/) to quantitatively estimate the effectiveness of preventative and interventive measures to prevent and battle COVID-19 outbreaks for specific populations.

**Materials and Methods:** Our simulator simulates the entire course of infection and transmission of virus among individuals in heterogeneous populations, subject to operations and influences such as quarantine, testing, social distancing, and community infection. It provides command-line and Jupyter notebook interfaces and a plugin system for user-defined operations.

**Results:** The simulator provides quantitative estimates for COVID-19 outbreaks in a variety of scenarios and assists the development of public health policies, risk-reduction operations, and emergency response plans.

**Discussion:** Our simulator is powerful, flexible, and customizable, although successful applications require realistic estimation and robustness analysis of population-specific parameters.

**Conclusion:** Risk assessment and continuity planning for COVID-19 outbreaks are crucial for the continued operation of many organizations. Our simulator will be continuously expanded to meet this need.

## Background and Significance

The worldwide COVID-19 pandemic has made everyone susceptible to life-threatening outcomes of the SARS-CoV-2 infection and made previously normal social and economic actions potentially hazardous. From healthcare facilities and government agencies to factories and businesses, organizations need tools to estimate risks of various operations to minimize risks of an outbreak, and develop emergency plans to contain the spread of the virus in the event of an outbreak.

While numerous epidemiological simulators have been developed, none are designed specifically for risk management and continuity planning, focusing on specific populations. In response to requests from various industries, hospitals, and government agencies, we developed a population-based simulator to study the outbreak of COVID-19 at an individual level and subject to various preventative measures and post-outbreak operations. We have been particularly responding to requests from actors, the shipping industry and schools for which a specified population with limited ingress is permitted, but for which controlling transmission is a critical requirement. Statistics summarized from the simulations have been used to assist the development of public health policies, risk-reduction operations, and emergency response plans for various environments.

## Materials and Methods

The basic assumption of the simulator is a population in which everyone is susceptible although certain groups could be more infectious or more susceptible than others. One or more virus carriers are introduced to the population at the beginning of or during the simulation (e.g. after a certain period of quarantine) and can potentially transmit the virus to others. If anyone shows any symptom, or is detected to be carrying the virus through testing, he or she can be removed, quarantined, or be kept in the population (e.g. in cases of ignored mild symptoms or incomplete quarantine). A simulation stops after pre-specified time, or when no one gets infected or shows symptoms or when everyone in the population is infected.

The core of the simulator is a statistical model that models the entire course of infection of infected individuals. The current model, which is likely to be updated with our deepening understanding of the epidemiology of COVID-19, models the incubation period of infected individuals with a lognormal distribution (1). Because it was reported that most pre-symptomatic transmission exposure occurred 1– 3 days before a person developed symptoms and that viral loads are already at peak and declining at the onset of symptoms (2), we designed an infectiousness model with transmissibility intensities that peak before the onset of symptoms and decline to a safe level 7 days later (Fig 1A).

**Figure 1:**
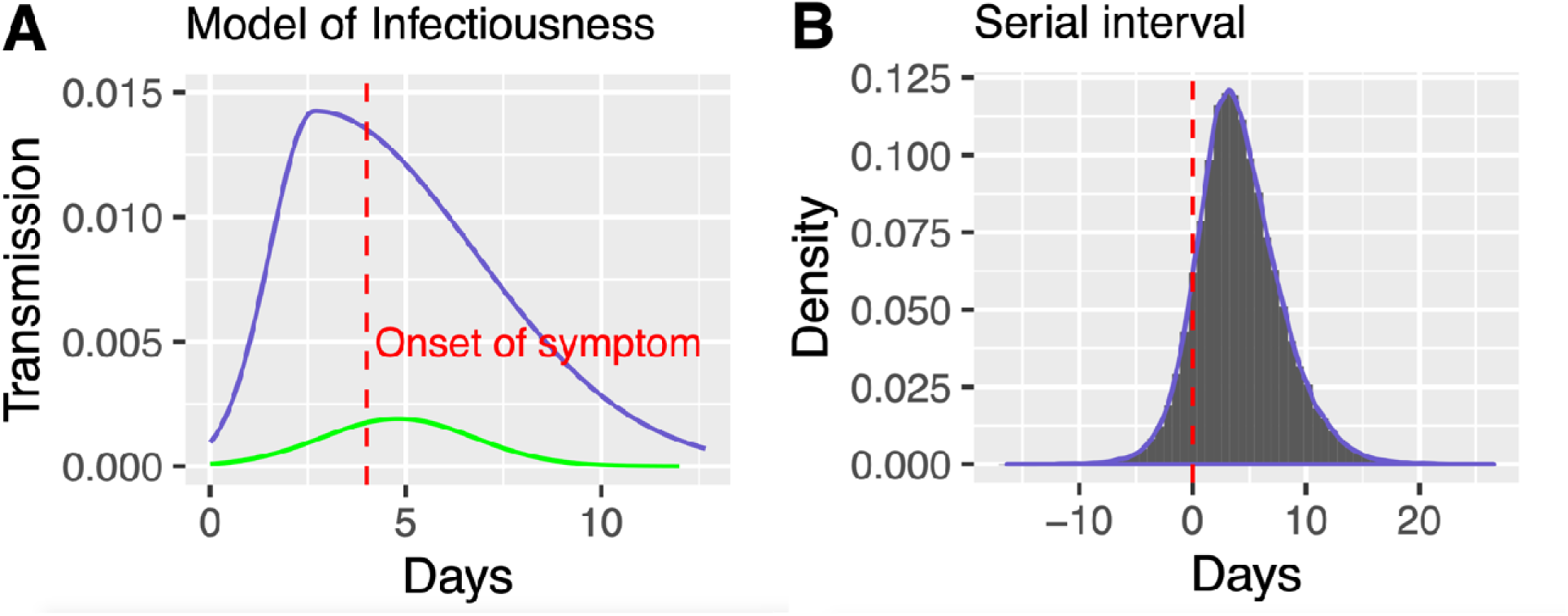
a) Probability of transmission per day for a symptomatic case with an incubation period of 4 days (purple line), and for an asymptomatic case (green line). b) Distribution of serial intervals of 10,000 simulated infector-infected individual pairs

Carriers who stay asymptomatic during their entire disease course tend to have lower viral loads than symptomatic carriers (3) and are less infectious, yet they could be a major factor for how wide-spread COVID-19 infection is. We allow asymptomatic cases to have default production number (R0) ranging from 0.28 to 0.56, which is one-fifth the R0 of symptomatic cases, with transmissibility probabilities that peak at 4.8 days and stop after 10 days. Because the exact proportion of asymptomatic transmissions is currently unknown and expected to vary from population to population, we allow asymptomatic transmission to be set as a constant or vary along a distribution, with default values set to a normal distribution centered at 25%, with a 95% confidence interval (CI) from 10% to 40% (4). We note that individuals in a population could have different model parameters and all modeling parameters, including distributions of some of the models, could be customized so our simulator is capable of simulating heterogeneous populations and populations with particular virus transmission models.

We validated our default model with extensive simulations and compared characteristics of simulated datasets, such as generation time, serial intervals, and proportions of asymptomatic, pre-symptomatic, and symptomatic infections, with other reports and models. For example, Fig. 1B shows the distribution of serial intervals of 10,000 simulations. The distribution follows a normal distribution with about 12% of pairs showing negative serial intervals (the infected individuals show symptoms before the infector), which is consistent with the observed data (5). The proportions of asymptomatic, pre-symptomatic, and symptomatic transmissions are 6%, 48%, and 44%, respectively, with default parameters (4).

This simulator is provided as a Python package with a command-line interface. It utilizes an event-driven model that changes the states of individuals in a population. A number of plugins are provided to simulate particular ways to introduce the SARS-CoV-2 virus to the simulated population (e.g. community infection), ways to detect affected individuals (e.g. PCR-based tests), to handle identified cases (e.g. quarantine), and to change model parameters dynamically (e.g. adjust of production number to model increased social distancing). The simulator can run tens of thousands of simulations on multiple processors in a few minutes, although the actual performance of the simulator depends on choice of parameters, especially the population size.

The simulator tracks events such as quarantine, reintegration, infection, onset of symptom, and removal of symptomatic cases and records outputs, such as time of events and number of infected individuals for each infector. Numerous summary statistics are calculated from replicated simulations to answer critical questions needed for risk assessment and continuity planning.

As examples of applications of the simulator, and sometimes as a higher-level interface, we provide simplified versions of the simulation studies we have performed as Jupyter notebooks, with narratives, simulation commands, and analysis of results. The entire analyses could be repeated by executing the notebooks, and some practitioners may accept default parameters, allowing the generation of prespecified reports for different model assumptions. We provide docker containers to allow the execution of our simulator and notebooks without the need to install relevant tools. Detailed documentations and instructions on the installation and execution of the simulator are provided on the project homepage.

## Results

With a flexible and customizable plugin system, the COVID-19 Outbreak Simulator is capable of simulating very complex scenarios in heterogenous populations. The applications section of our homepage lists examples of some applications that we have performed for various real-world projects and we expect the list to grow over time.

As a basic example, we simulated a scenario with an infected individual entering an enclosed environment without quarantine. We assumed that from 10% to 40% of infected individuals never showing any symptoms, and those who show symptoms will be removed from the population promptly. Under those parameters and assuming one infected individual enters the environment at the start of the simulation, in 18.2% of all cases, the carrier will not infect anyone or show any symptoms. In the remainder of cases, an outbreak will happen, and unless the carrier shows symptoms on the first day, there is a 47.3% probability the virus has already been transmitted to at least one other person by the time of detection, so removal of symptomatic cases is insufficient to stop the outbreak. The probability that a second symptomatic case happens remains high until 1 week after the removal of the first case. Fig. 2a illustrates the duration of outbreak versus the remaining population size in this scenario, showing that a large percentage of outbreaks stop on the first day when the carrier, most likely asymptomatic carriers, are not expected to infect anyone else. Based on the high probabilities of an outbreak under this scenario better approaches to risk management require quarantine prior to the introduction of infected individual to the population. A 7-day quarantine would avoid 82.1%, and a 14-day quarantine would avoid 99.3% of potential outbreaks (Fig. 2b).

**Figure 2:**
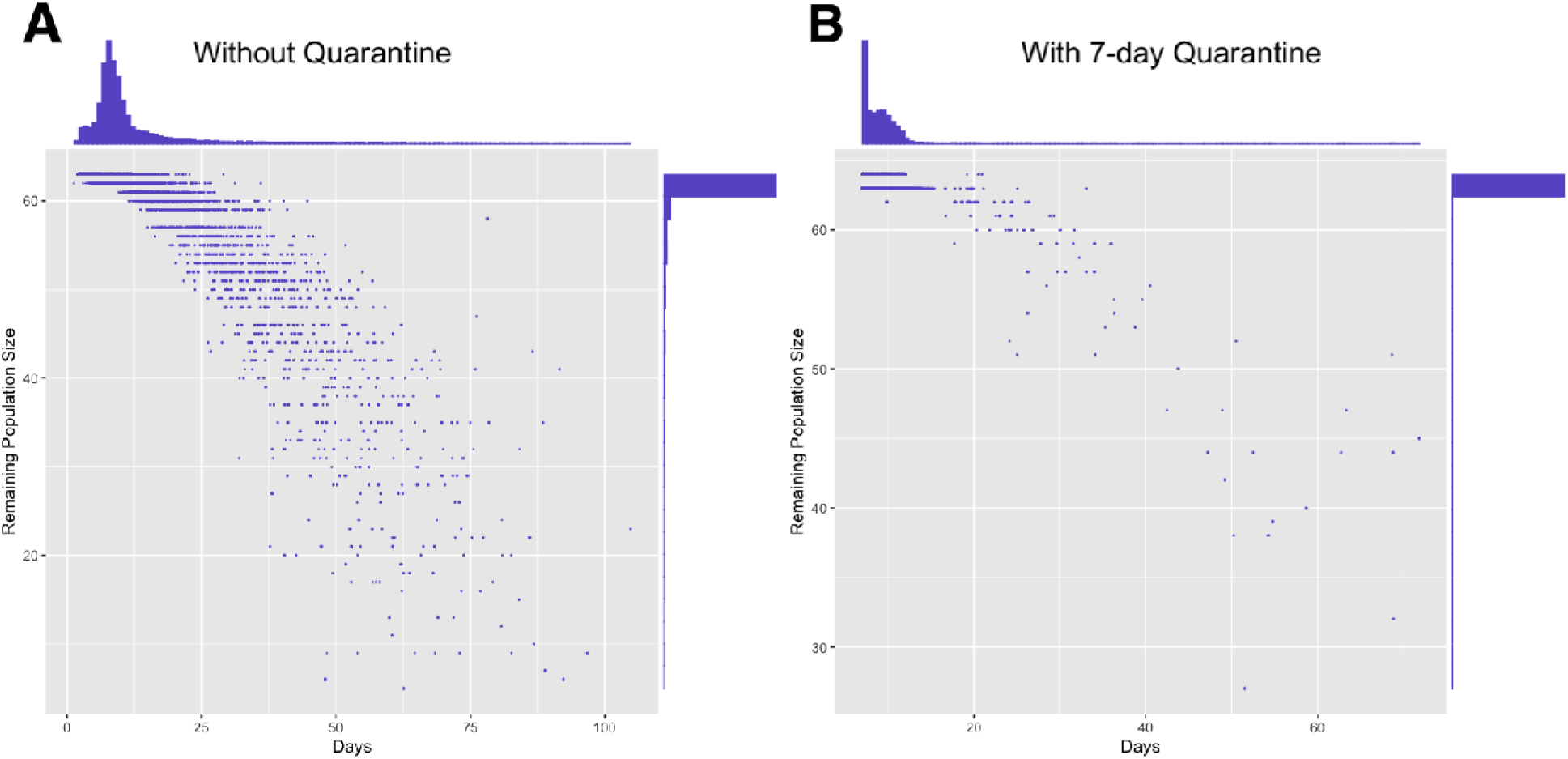
Duration vs remaining population size for 10,000 simulated outbreaks. A) The virus carrier was introduced to the population as long as he or she did not show any symptom. B) The virus carrier was introduced to the population after 7-day quarantine.

As a second example, we simulated a lab environment in which lab members are subject to infection from community and family members they interact at home. To ensure safety of the lab environment, the manager would like to test everyone periodically so that infected lab members, even if they are asymptomatic or presymptomatic could be identified and quarantined before infecting others. The simulations assumed that everyone has an equal probability of getting infected (p=0.0022), and PCR tests are performed at every 3, 7, or 14 days, or not performed at all. As we can see from Table 1, despite the seeming low probability of community infection, almost all labs have at least one case of SARS-CoV-2 infection after 90-days and the percentage of uninfected labs decreases quickly with increasing lab size. Periodical PCR-tests can successfully reduce within-lab infections but the manager must strike a balance between cost and benefit to determine frequency of testing. The simulator can further simulate accuracy of tests and delayed return of test results to better mimic real-world scenarios.

**Table 1:** Effectiveness of periodic PCR-based tests in reducing within-lab infections. All simulations assumed a daily probability of community infection of 0.0022. Results for each scenario are based on 10,000 replicate simulations.

| Lab Size | Test frequency | Average Lab Size after 90 days | Proportion of Uninfected Labs | Average Number of Community Infection | Average Number of Within-Lab Infection |
| --- | --- | --- | --- | --- | --- |
| 10 | 3 | 5.9 | 1.14% | 3.6 | 0.5 |
| 10 | 7 | 5.2 | 1.14% | 3.4 | 1.5 |
| 10 | 14 | 4.5 | 1.15% | 3.1 | 2.4 |
| 10 | No test | 3.7 | 1.01% | 2.9 | 3.5 |
| 20 | 3 | 11.8 | 0.02% | 7.1 | 1.2 |
| 20 | 7 | 10.1 | 0.03% | 6.6 | 3.3 |
| 20 | 14 | 8.3 | 0.00% | 6.1 | 5.6 |
| 20 | No test | 6.2 | 0.00% | 5.3 | 8.5 |
| 30 | 3 | 17.6 | 0.00% | 10.6 | 1.9 |
| 30 | 7 | 15 | 0.00% | 9.9 | 5.2 |
| 30 | 14 | 12 | 0.00% | 9 | 9 |
| 30 | No test | 8.5 | 0.00% | 7.7 | 13.8 |

## Discussion

Simulations of the outbreaks of COVID-19, especially in complex scenarios, require the specification of a large number of parameters, many of which are difficult to determine. Despite best efforts to provide parameters with most likely values, the simulated scenarios might deviate significantly from the reality, compromising the validity of predictions. In addition to constant update of the baseline model according to most recent discoveries about the epidemiology of COVID-19 outbreaks, we allow the specification of distributions, instead of fixed values for many parameters to account for variations of models. We also perform a large number of simulations with different parameters to obtain predictions associated with a parameter space and provide results under best and worst scenarios. During this process we were able to not only assess the robustness of our predictions, but also identify parameters that have the most impact on outcomes (e.g. severity of the outbreak), which by itself were valuable for policy making purposes.

A limitation of the simulator is that it does not simulate physical locations, so the transmission of virus is performed by probability, not by actual physical vicinity or contact. This design avoids a large number of model assumptions such as population boundaries and individual movement patterns and allows much more efficient simulations of large populations. It however makes it currently infeasible to simulate scenarios such as transmission through common traffic hubs such as schools and supermarkets. Despite this limitation, the current version of the software could be useful for studying the behavior of outbreaks affecting many environments in which individuals are well mixed. We have been asked to provide guidance for a wide variety of industries, including shipping, mining, actors and laboratory environments for which a limited population may interact regularly. Our simulator is effective for identifying how risk can be managed and what kinds of mitigation provide acceptable controls for these situations. We are expanding the simulator, mostly by adding more plugins, to accommodate more scenarios. The software is available through GitHub and further modifications can be made by us or other users.

## Conclusion

The COVID19 Outbreak Simulator simulates the spread of the SARS-CoV-2 virus in environments with heterogeneous but well-mixed populations, subject to operations such as quarantine, testing, social distancing, and influences such as community infection and incomplete detection. It provides quantitative measures to assess risks of an outbreak associated with various operations, and assist the development of emergency plans in response to an outbreak. The simulator has been successfully used for applications in industries, schools and government agencies. Although simulating and analyzing particular scenarios with realistic assumptions can be challenging, the growing number of notebooks with simulated scenarios for particular environments, potentially contributed by our users, would make the simulator easier to use and provide guidance to epidemiologists and policy makers without the need to execute the simulator.

## Data Availability

All source code and documentation of this project is publicly available at https://github.com/ictr/covid19-outbreak-simulator

## Acknowledgements

The authors appreciated feedbacks from users of this simulator and Benjamin Wu, a summer student for his contributed to the data analysis part of the project.

## Availability of data and materials

The COVID-19 Outbreak Simulator is released through Python Package Index under the name covid19-outbreak-simulator. All data generated and/or analyzed during the current study, in particular applications in the format of Jupyter notebooks are publicly available through https://github.com/ictr/covid19-outbreak-simulator/.

## Authors’ contributions

CIA conceptualized the idea of the simulator and applied it to real-world applications. BP implemented the simulator, performed all the simulations and data analyses, and drafted the manuscript. Both CIA and BP reviewed and approved the manuscript.

